# High burden of non-adherence to blood pressure-lowering medications: meta-analysis of data from over 34,000 adults with hypertension in Sub-Saharan Africa

**DOI:** 10.1101/2024.05.28.24308082

**Authors:** Leopold Ndemnge Aminde, Valirie Ndip Agbor, Noah Takah Fongwen, Calypse Ngwasiri, Clovis Nkoke, Miriam Nji, Anastase Dzudie, Aletta E. Schutte

**Author notes:** **Correspondence:** Leopold N. Aminde, MD PhD FFPH. Public Health & Economics Modelling Group, School of Medicine & Dentistry, Griffith University, Gold Coast, QLD 4222, Australia.

## Abstract

**Introduction:** Non-adherence to blood pressure (BP)-lowering medication is a strong predictor of poor BP control. Sub-Saharan Africa (SSA) has extremely low BP control rates (∼10%), but it is unclear what the burden of medication non-adherence among Africans with hypertension is. This systematic review estimated the prevalence and determinants of non-adherence to BP-lowering medications in SSA.

**Methods:** Multiple databases were searched from inception to 6 December 2023. Two reviewers performed independent screening, extraction, and quality assessment of studies. We pooled the prevalence estimates using random effects meta-analyses and summarized the determinants using a narrative synthesis.

**Results:** From the 1,307 records identified, we included 95 studies published between 1995 and 2023. The overall prevalence of non-adherence to BP-lowering medication among 34,102 people treated for hypertension in 27 countries was 43.5% (95% confidence interval 39.4 to 47.6; I^2^ = 98.3%). There was no change in the prevalence of non-adherence over time. The burden of non-adherence varied by measurement method (p = 0.028) and by median age (38.8%, > 57 years vs. 47.9%, ≤ 57 years; p = 0.015). Socioeconomic and patient-related factors were the most frequent factors that influenced medication adherence. Active patient participation in management, accurate perceptions, and knowledge of hypertension and its treatment predicted good medication adherence, whereas high pill burden, medication cost, side effects, and comorbidities predicted poor adherence.

**Conclusions:** With the African population projected to increase from 1.4 to ∼2.5 billion by 2050, targeted strategies are urgently needed to optimise medication adherence in people with hypertension in SSA.

## Introduction

Hypertension is the leading risk factor for disease burden globally, affecting nearly 1.3 billion adults (1), and contributing to over 10.8 million deaths each year (2). The prevalence of hypertension in sub-Saharan (SSA) is high, affecting about one in three adults and one in two people aged 50 years and over (3–6). In SSA, over half of the women and two-thirds of the men with hypertension remain undiagnosed. This high burden of hypertension contrasts with very low treatment and control rates. A recent global analysis found that only 22–29% of adults diagnosed with hypertension in SSA are treated; of these, only 9% of men and 13% of women have their blood pressure (BP) controlled (1). Pharmacotherapy and lifestyle modification are the mainstay of BP control to prevent vascular complications including stroke, ischemic heart disease, chronic kidney disease, and dementia (3).

The World Health Organization (WHO) defines adherence as the extent to which a person’s behaviour— taking medication, following a diet, or executing lifestyle changes—corresponds with agreed recommendations from a healthcare provider (7). Non-adherence to medication has been identified as a major predictor of poorly controlled hypertension. Compared to people who are adherent to BP-lowering medication, those who are non-adherent are twice as likely to have sub-optimal BP control and to develop complications of hypertension (8). Understanding the extent of non-adherence and determinants is critical to the development of appropriate interventions to prevent the health and economic consequences for affected persons and health systems. Prior reviews exploring adherence to BP-lowering medication have been limited by either narrow inclusion time periods (9, 10), restricted to one adherence measurement method (9, 11), lacked quality appraisal of the evidence (9), did not assess determinants of non-adherence (12), or had a broad geographical focus with limited representation of SSA (8, 13).

Given the high burden of hypertension, low rates of BP control in SSA, and the existing evidence gaps, this systematic review and meta-analysis sought to answer the following questions: 1) What is the contemporary burden of non-adherence to BP-lowering medication among people with hypertension in SSA, and has it changed over time? and 2) What are the determinants of (non-)adherence to BP-lowering medication in SSA?

## Methods

### Search strategy and selection criteria

The review protocol is registered in the PROSPERO database (CRD42017079838) and published (14). This review was reported according to the Preferred Reporting Items for Systematic reviews and Meta-analysis (PRISMA 2020) guidelines (15). We searched MEDLINE via Ovid, SCOPUS, Web of Science, African Journals Online (AJOL) databases and grey literature through Google Scholar to identify all relevant articles from the inception of databases until 6 December 2023. Search terms related to BP-lowering medication and adherence were combined with relevant Boolean operators and a validated African search filter (16) to improve the sensitivity and precision of the search. No language restrictions were applied. See **Tables S1-S4** for the detailed search strategies.

We included primary observational studies reporting the prevalence of or factors associated with (non-) adherence to BP-lowering medication among adults with hypertension residing in SSA. We excluded studies conducted out of SSA, studies that selected participants based on their medication adherence status, and those with incomplete data or lacking relevant data to compute the prevalence of non-adherence. We excluded studies with less than 30 participants, letters to the editor, commentaries, reports without primary data, or explicit descriptions of the methods. Duplicate studies based on the same data were excluded and only the most comprehensive report was retained.

### Screening and data extraction

Following the database search, we uploaded all records to the Rayyan systematic review software (17) and removed all duplicates. Two investigators (LNA and FNT) independently screened titles and abstracts. Full texts of potentially eligible studies were assessed for final inclusion (LNA and VNA), and data was independently extracted by two reviewers (LNA, VNA, FNT, MAN, CAN, and CN). We resolved any disagreements by consensus discussion. The following data were extracted from individual studies: the last name of the first author and year of publication, year(s) of recruitment, the country, study design, study setting (rural vs urban) and study type (community-based vs. hospital-based), method or tool used to define medication adherence, approach to data collection (interview-administered vs. self-administered), sample size, total number of patients non-adherent to BP-lowering medication, mean or median age, and the male proportion. For multinational studies, we extracted the combined data first and, where possible, the disaggregated (country-specific) data. For studies reporting on factors associated with medication adherence, we extracted measures of association and confidence intervals for variables based on multivariable regression analyses.

### Risk of bias assessment

We used the risk of bias tool developed by Hoy *et al*. (18) to evaluate the methodological quality of prevalence studies. The tool assesses selection/sampling methods, response rate, outcome measurement and validity of reporting. We used the Newcastle-Ottawa Scale (NOS) to assess cohort studies (19), which includes eight items spread across three domains: selection of participants, comparability, and outcome assessment. Two reviewers (LNA and CAN) independently assessed the quality of studies and any disagreements resolved by consensus.

### Data analysis

A random effects meta-analysis using the DerSimonian-Laird method was used to estimate the pooled prevalence of non-adherence to BP-lowering medication. We used the Freeman-Tukey double arcsine transformation to stabilize the variance before meta-analysis (20). We assessed between-study heterogeneity using Cochran’s Q test and the I^2^ statistic. I^2^ values < 30%, 30–49%, 50–74% and ≥75% represented low, moderate, substantial, and considerable variation in study-specific estimates due to heterogeneity (21). We performed multiple subgroup analyses to investigate potential sources of heterogeneity. The Q test based on the analysis of variance was used to compare between subgroups. For studies reporting high, medium, and low adherence levels, as opposed to adherent vs. nonadherent, we defined non-adherence as having low adherence levels in the main analysis. We performed sensitivity analyses in which we classified medium adherence as non-adherent. Additionally, we estimated the prevalence of non-adherence using only studies with low risk of bias. We conducted an influence analysis to investigate the impact of individual studies on the pooled estimate.

To examine for small study effects, we used the Doi plot and the Luis-Furuya Kanamori (LFK) index. Compared to the traditional funnel plot and Egger’s regression test, these measures have been shown to have better visual representation of asymmetry and diagnostic accuracy in detecting small study effects. An LFK index of zero represents complete symmetry in the Doi plot, while any values beyond ± 1 are consistent with plot asymmetry (22, 23). All statistical analyses were conducted in R Studio (R version 4.2.3) using the ‘meta’ and ‘metasens’ packages.

## Results

### Overview of included studies

The search identified 1,315 records. After removing duplicates and screening titles and abstracts, we assessed 158 full texts for eligibility. We included 95 studies comprising 34,102 adults with hypertension in the systematic review and meta-analysis (**Figure 1**). There were two multi-country studies, one with twelve countries (24) and the other with two countries (25). Overall, most of the studies were conducted in Western SSA countries (n = 50, 47.2%), including Nigeria (n = 29), Ghana (n = 11), Côte d’Ivoire (n = 4), Togo (n = 2), and Benin, Guinea, Mauritania, Senegal with one study each. The second most represented region was Eastern SSA (n = 34, 32.1%), including Ethiopia (n = 23), Kenya (n = 3), Eritrea and Tanzania with 2 studies each, and Malawi, Mozambique, Seychelles, and Uganda with one study each. Next was Southern SSA (n = 13, 12.3%), with studies conducted in South Africa (n = 5), Zimbabwe (n = 3), Namibia (n = 2) and one each in Botswana, Lesotho, and Zambia. Finally, studies from Central SSA (n = 9, 8.5%) included Cameroon (n = 4), the Democratic Republic of Congo (n = 2), Congo (n = 2) and one in Gabon.

**Figure 1:**
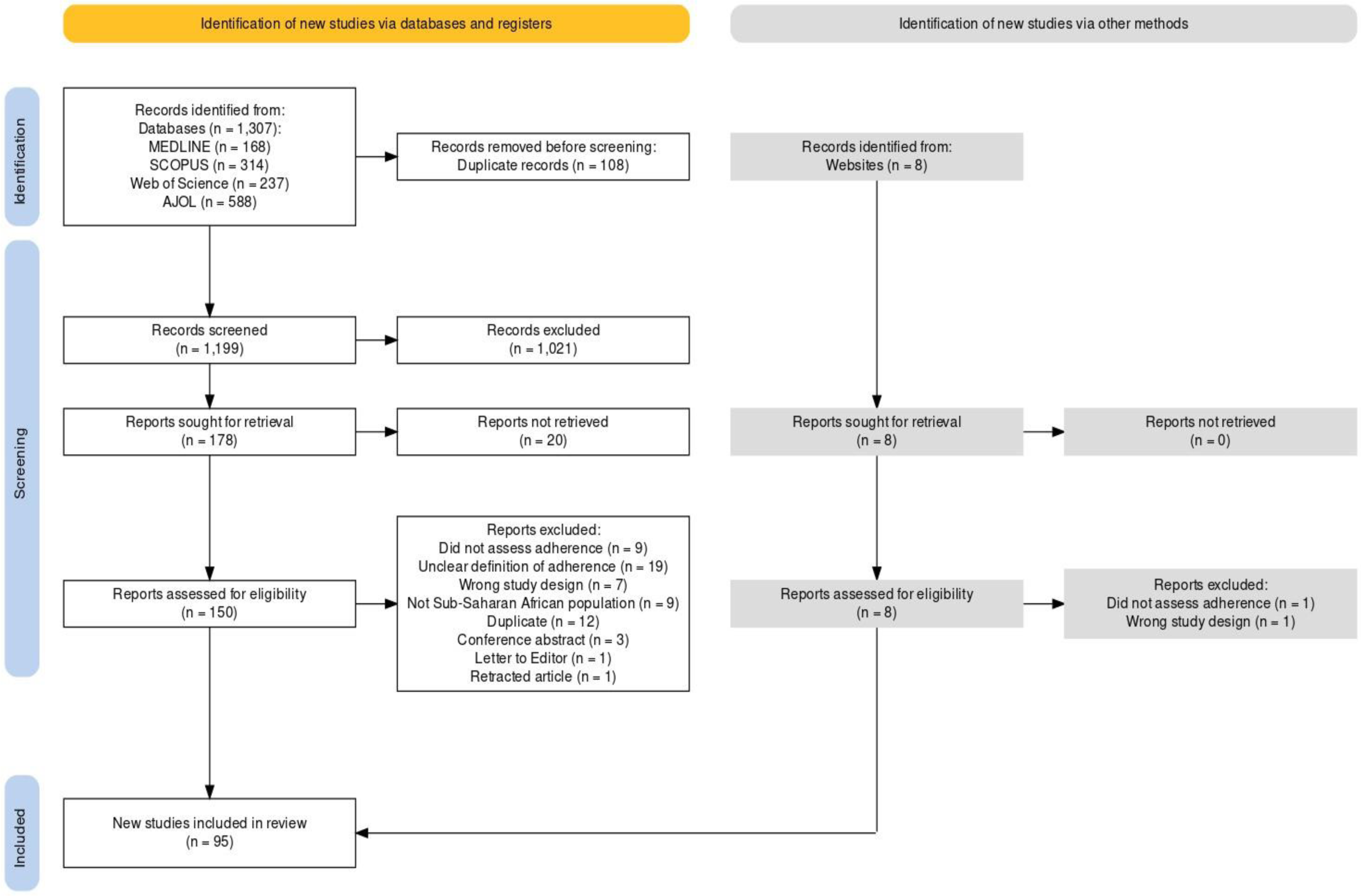
PRISMA flow diagram depicting the selection of studies for inclusion in the review.

The most frequently used tool to measure adherence was the eight-item Morisky Medication Adherence Scale (MMAS-8) (n = 25, 26.3%) (24–48). In 21 studies (22.1%), the authors used the proportion of days medication was taken (PDMT ≥ 80%) per week (49–69). The next most frequently used tool was the Hill-Bone Compliance Questionnaire (n = 11, 11.6%) (70–80). The pill count method was used in six studies (6.3%) (81–86), and only one study used biological assays (87). Most studies were cross-sectional (n = 89, 93.7%), whereas six were cohort studies (6.3%). Forty-eight (50.5%) studies were conducted in urban settings, five (5.3%) in rural, and forty-two (44.2%) in both urban and rural settings. Almost all the included studies were hospital-based (93.7%). About three-quarters (n = 67, 70.5%) of studies were rated as having a low risk of bias, whereas the remaining studies (n = 28, 29.5%) had a moderate risk of bias. (**Table 1** and **Tables S5-S7**).

**Table 1:**
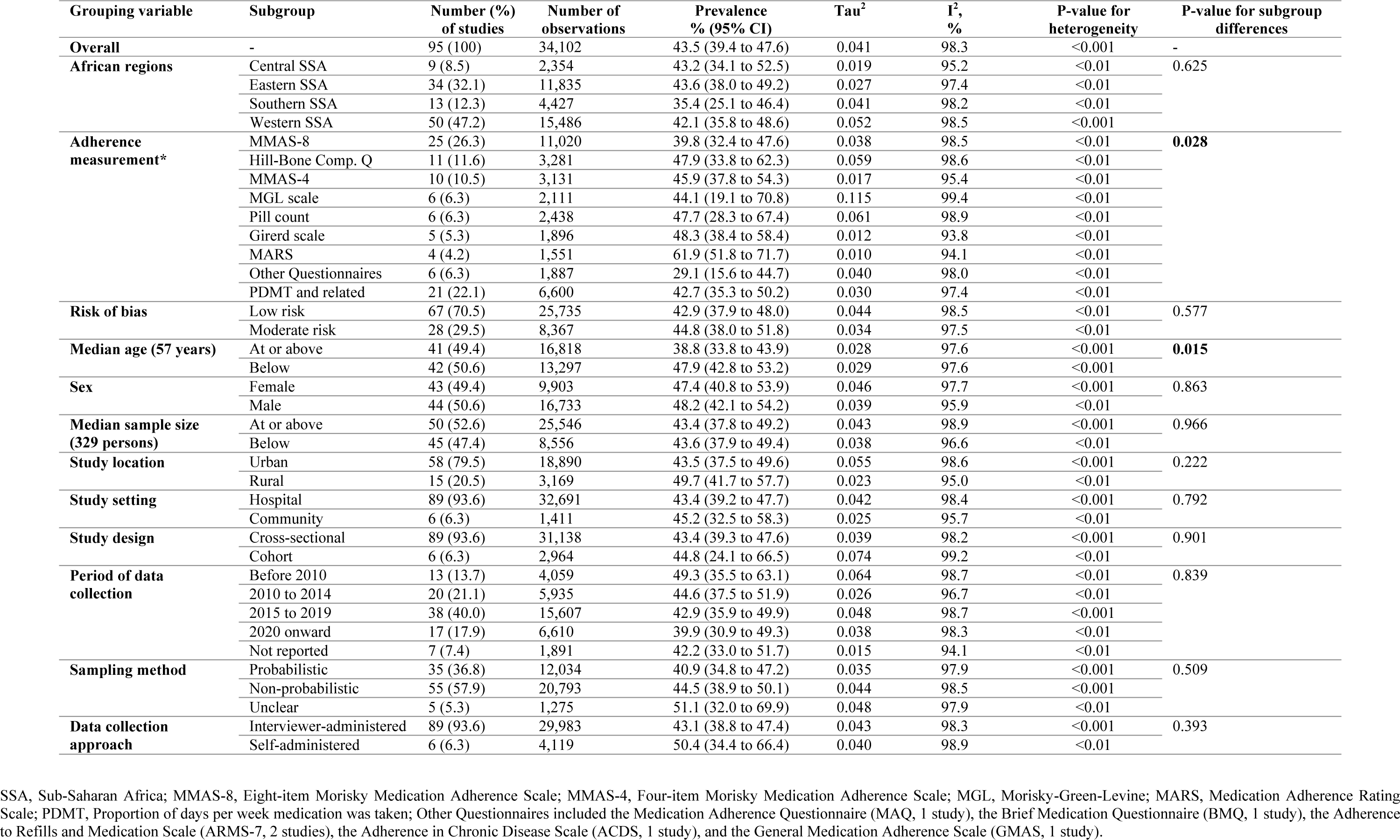
Sub-group analysis exploring differences in prevalence of non-adherence to BP-lowering medication in Sub-Saharan Africa.

### Burden of non-adherence to BP-lowering medication

#### Overall

The prevalence of non-adherence to BP-lowering medication from individual studies ranged from 3.5% (26) to 92.9% (69). The pooled prevalence of non-adherence across all studies was 43.5% (95% CI: 39.4 – 47.6), with substantial between-study heterogeneity (I^2^ = 98.3%, *p* < 0.0001) (**Figure 2**). Meta-regression analysis showed no change in the prevalence of non-adherence over time (coefficient: −0.004, 95%CI: −0.011 to 0.004), (**Figures S1 and S2**). In sensitivity analysis, the prevalence of non-adherence was similar to the main analysis estimate when considering only studies with low risk of bias (**Figure S3**). However, classifying people in the medium adherence category (n=19 studies) as being non-adherent to their medication resulted in a higher overall prevalence of non-adherence (51.3%, 95%CI: 46.1 – 56.6; I^2^ = 99%) (**Figure S4**).

#### Sub-group analysis

The burden of non-adherence to BP-lowering medication varied by age (*p* = 0.014), with younger participants (< median age 57 years) having greater non-adherence (47.9%; I^2^ = 97.6%) compared to older participants (≥ 57 years) (38.8%; I^2^ = 97.6%) (**Table 1** and **Figure S5**). Non-adherence also varied by measurement tool (*p* = 0.028). The prevalence of non-adherence was 39.8% (25 studies, I^2^ = 98.5%) for the most frequently used tool (MMAS-8), lowest (29.7%) among studies using other questionnaires (7 studies, I^2^ = 98.0%) and highest (61.9%) in studies using the Medication Adherence Rating Scale (4 studies, I^2^ = 94.1%) (**Table 1** and **Figure S6**). There was no sex difference in non-adherence to BP-lowering medication (47.4%, 43 studies with 9,903 women vs. 48.2%, 44 studies with 16,733 men, *p* = 0.855). There was no statistical difference in the prevalence of non-adherence between Southern SSA (35.4%, 13 studies, I^2^ = 98.2%) compared to Western SSA (42.1%, 50 studies, I^2^ = 98.5%), Central SSA (43.2%, 9 studies, I^2^ = 95.2%) and Eastern SSA (43.6%, 34 studies, I^2^ = 97.4%) (*p* = 0.613). Similarly, the prevalence of non-adherence did not differ by location (49.7% for rural vs. 43.5% for urban; *p* = 0.233), study settings, study design, sample size, sampling method, period of data collection or data collection approach (**Table 1** and **Figures S7–S16**).

### Influence analysis and publication bias

The influence analysis found no evidence that any individual study-specific estimate significantly influenced the pooled estimate (***Figure S17***). Additionally, we found no evidence of small study effects as the Doi plot was visually symmetrical, and confirmed with the LFK statistic (LFK index = 0.56) (**Figure S18**).

### Factors associated with medication non-adherence

**Figure 3** and **Table S8** present the details of the predictors of medication non-adherence, which are summarised below.

**Figure 3:**
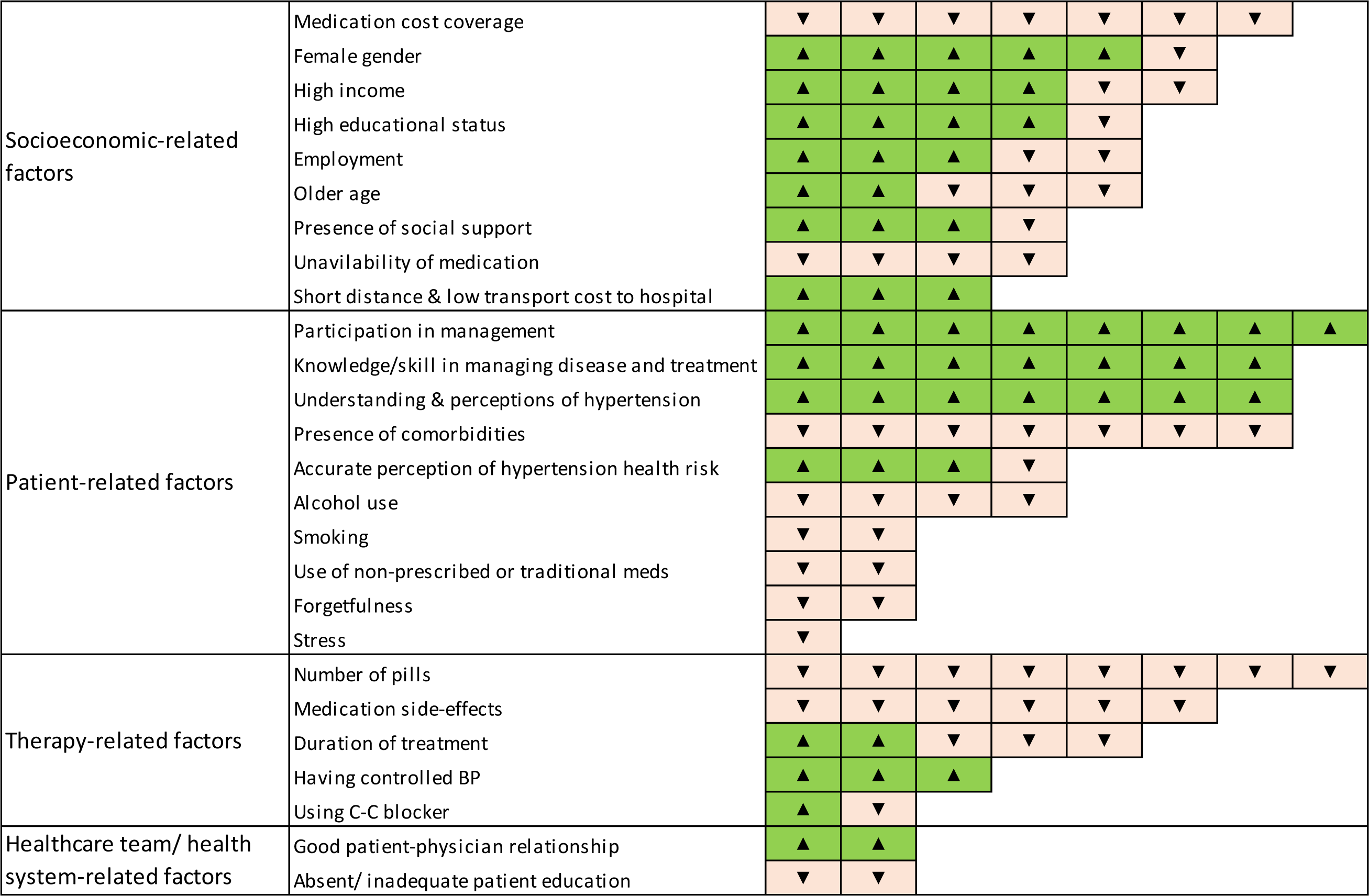
Factors associated with medication (non-)adherence in sub-Saharan Africa classified according to the World Health Organization dimensions for adherence to medication in people with hypertension.

#### Demographic and socioeconomic factors

Age, sex, and area: All but one (69) of six studies found that being male was associated with non-adherence to medication (88–92). Additionally, three out of five studies showed younger adults (generally < 40 years) were more likely to be non-adherent to medication compared to older adults (48, 84, 93), but two studies found the contrary (89, 94). One study reported that residing in rural areas was associated with non-adherence (94).

Education: All five studies showed that lower levels of education were associated with non-adherence to medication (41, 67, 82, 86, 95).

Employment and income: Being unemployed was associated with poor medication adherence in four studies (32, 71, 90–92), whereas three authors found the opposite relationship (32, 43, 67). Additionally, three (24, 67, 76) out of five studies reported that higher income/wealth status was associated with good medication adherence, whereas the converse was found in two studies (71, 93).

Social support: All but one (71) of four studies (32, 41, 71, 75) noted that having good family/friend support was associated with good medication adherence, with odds ratios ranging from two (32) to over five-fold (75).

Distance from health facility: Two studies showed that people experiencing high cost or lack of transport had over three times the odds of being non-adherent to medication (72, 92); individuals with shorter distances to the hospital had a two-fold higher odds of being adherent (aOR: 2.02, 1.19 to 3.43) (88).

Medication cost and availability: Unavailability of medication was associated with non-adherence to medication (71, 81, 95, 96), with the odds ranging from 1.7 (95%CI: 1.1 to 3.3) in a study in Tanzania (81) to 5.4 (95%CI: 1.76 to 16.85) in Namibia (71). All seven studies investigating cost coverage for medication found that compared to having free medication or funded by others/insurance/government schemes, self-funded or out-of-pocket costs were associated with non-adherence to medication (34, 36, 41, 46, 48, 81, 95).

#### Patient-related factors

Active participation in management: All eight studies found that patients who actively participated in the management of their BP through regular attendance to clinic appointments (71, 75, 91), health talks (92), having good self-efficacy (82, 90) and not stopping medication when asymptomatic (70, 81) were more likely to have good medication adherence.

Understanding and perceptions about hypertension: Patients with a good understanding of hypertension were three to nine times as likely to be adherent to their medication compared to those who had a poor understanding of the condition (27, 34, 45, 88–90, 92).

Knowledge and skill in managing symptoms and treatment: Patients with adequate knowledge and skill had 3 to 13-fold higher odds of being adherent to medication compared to their counterparts (30, 37, 70, 87, 88, 94, 95, 97).

Perception of the health risk related to hypertension: All but one (27) of four studies found that people with accurate perceptions had higher odds of being adherent to their medication (32, 96, 97).

Comorbidities: In six studies, patients with one or more comorbidities were over twice as likely to be non-adherent to their medication (36, 43, 61, 84, 88, 95). One study reported that those without a comorbid condition were about four times as likely to be adherent to their medication (34).

Alcohol use and smoking: All studies evaluating the impact of alcohol use (27, 36, 37, 70) and smoking (61, 70) found that both behaviours were associated with low or non-adherence to medication.

Stress and forgetfulness: Patients who reported being stressed were twice as likely to be non-adherent to medication, whereas being forgetful was associated with about 8-fold higher odds of non-adherence to medication (45, 46).

Others: Persons using non-prescribed medication (96) or traditional medications (24) were twice as likely to be non-adherent to their BP-lowering medication. Similarly, being physical inactive (aOR: 1.63, 1.04 to 2.55) was associated with non-adherence to medication (43).

#### Therapy-related factors

Pill burden: Eight studies reported that high pill burden was associated with non-adherence to medication (36, 46, 84, 86, 89, 93, 94, 98), with poorer adherence in people taking two or more BP-lowering pills (36, 46, 89, 94).

Side-effects: All six studies showed that those who experienced side effects (such as dizziness (72), palpitation (70)) were two to seven times more likely to be non-adherent to their medication (37, 46, 70, 72, 82, 96).

Duration of diagnosis and treatment: Three studies showed that people who had been diagnosed or on treatment for longer than one (37), five (98) or ten years (93) were more likely to be non-adherent, while two studies showed that being on treatment for three or more years was associated with good medication adherence (34, 76).

BP control and class of BP-lowering medication: All three studies exploring the relation with BP control found that people with well-controlled BPs were two to three times more likely to be adherent to their medication (61, 88, 96). Two studies found a significant relationship between the use of calcium-channel blockers and medication adherence, although both had differing conclusions (24, 37).

#### Healthcare team/health system factors

Good patient-doctor relationships: This was associated with up to four-fold higher odds of adherence (34), whereas a poor relationship was associated with non-adherence to medication (95).

Health education: Two studies showed that patients who did not receive health education or counselling when attending health facilities were twice as likely to be non-adherent to their medication (41, 96).

## Discussion

This systematic review provides a comprehensive synthesis of the burden of non-adherence to BP-lowering medication among adults with hypertension in SSA. At least two out of every five adults on treatment for hypertension in SSA were non-adherent to their medication. Non-adherence was higher in younger (mean age < 57 years) adults compared to older adults, and it varied by measurement method. Socioeconomic- and patient-related factors were the most frequently reported drivers of medication adherence in SSA. There was consistent evidence that active patient participation in management, accurate understanding and perceptions, and knowledge of hypertension and its treatment predicted good adherence to medication, while high pill burden (mostly more than two pills per day), medication cost, experiencing side effects, and having comorbidities predicted poor adherence.

The overall prevalence of non-adherence in this study was lower than the 62.5% and 65.9% reported by previous studies in Africa (9, 10). These reviews had very narrow periods for which data was included, ranging from seven to eleven years, as opposed to the present review, which pooled data from studies published over the last three decades. Nevertheless, the estimates of the present study are on par with findings from a global meta-analysis that found a prevalence of non-adherence—measured using questionnaires—of 43% in LMICs, which was higher than the 26% to 38% reported from high-income countries (8).

Non-adherence to BP-lowering medication was higher in studies with younger populations compared to those with older persons, consistent with reports from both LMICs and HICs (8, 13, 99). Older adults are more likely to have chronic conditions and to dedicate more time to their health and therapy. Furthermore, the use of adherence aids like pill boxes and calendars likely enhances their ability to adhere to medication (100). We also found variations in the pooled prevalence of non-adherence by the tool used in assessing medication adherence, consistent with previous reviews (8, 13). There is currently no gold standard for measuring adherence. Thus, the observed variation could reflect differences inherent to the respective tools (101, 102).

Socioeconomic and patient-related factors were the major drivers reported for non-adherence to BP-lowering medications. This is consistent with prior systematic reviews (10, 11, 13). However, we found conflicting results for age and employment status. While some studies showed that older age was associated with good adherence, three out of five studies reported an inverse relationship. The latter could be due to declining cognitive ability with ageing. Being employed is correlated with higher socioeconomic status and access to healthcare services, which could explain better adherence (13). In contrast, the association between employment and non-adherence could be due to patients prioritizing their job schedules. These mixed results likely indicate a complex relationship between age, employment, and medication adherence and potential differences in study settings.

Active patient participation and good knowledge of hypertension, and their treatment were the main patient-specific factors consistently associated with good medication adherence. Targeted interventions promoting these attributes should be a key focus in SSA. There is growing evidence elsewhere about the benefits of active patient involvement in decision-making about their health and adherence to therapy (100, 103, 104). These benefits include increased satisfaction, quality of life, and improved understanding of hypertension.

Comorbidities, experiencing medication side effects, and taking multiple pills were associated with medication non-adherence. Similar results were reported among older adults (11). People with multiple chronic conditions are likely to have a high pill burden and incur significant out-of-pocket health expenditures (105, 106). This is a particular challenge for patients in LMICs such as SSA with limited insurance or universal health coverage (107). There is strong evidence that most people with high BP require two medications to control their BP. Hence, simplifying treatments through single-pill combinations (SPC) is likely to address the problem of pill burden and adherence, with spin-offs like improved quality of life, reduced clinical inertia and lower healthcare costs (108–110).

### Implications of the findings

The population of Africa is projected to increase from 1.4 billion now to ∼2.5 billion by 2050, representing the fastest population growth worldwide (111). Our analysis shows the prevalence of non-adherence to BP-lowering medication has not changed over time. With one in three adults having hypertension (4), health systems in SSA can expect to manage ∼825 million people with hypertension. Assuming this pattern continues, this would approximate to over 350 million patients not adherent to their medication, and at increased risk of cardiovascular events. This represents a substantial challenge for already struggling health systems. Hence, in addition to prioritising primordial prevention, governments in SSA need to develop sustainable longterm strategies. For example, accelerating progress towards universal health coverage for the delivery of quality and equitable healthcare for all, and fostering team-based care to improve healthcare provider-patient ratios and relationships (112). Furthermore, with SPCs now on the WHO Essential Medicines List (113), stakeholders involved in BP control should adopt simple and practical treatment guidelines incorporating SPCs. Additionally, governments need to facilitate supply chains that enhance the availability of these SPCs for patients. Such strategies would contribute to improving adherence to therapy, BP control and CVD outcomes.

### Strengths and limitations

The review had some limitations. First, most of the included studies were hospital-based. Hence, the estimate of medication non-adherence may be biased by the ‘white coat adherence’ phenomenon, whereby patients are more adherent in the days leading up to and immediately after their clinic visit (114, 115). Consequently, the prevalence presented in this study may be an underestimation of the true prevalence of medication non-adherence in SSA. Second, over half of the included studies were from western SSA, with lower representations from central and southern SSA. Furthermore, there were comparatively fewer studies conducted in rural areas. Hence, the estimate from this review might not be generalizable to the whole of SSA. Third, there was substantial heterogeneity between the studies, which was not completely explained by the variables in the sub-group analyses. Furthermore, the prevalence of non-adherence varied based on the adherence measurement method. More research is needed to develop standardized cost-effective and pragmatic tools for assessing medication adherence for research and clinical practice in SSA. Despite these shortcomings, this systematic review provides a comprehensive synthesis of the evidence from mostly high-quality studies on the burden and determinants of non-adherence to BP-lowering medication in SSA.

## Conclusions

This systematic review showed a substantially high burden of non-adherence to BP-lowering medication among adults with hypertension in SSA; two out of every five adults treated for hypertension are not adherent to their therapy, and this has not changed over the last three decades. Targeted policies are needed to address the broad range of socioeconomic, health system, therapy, and patient-related factors driving this high burden of medication non-adherence in SSA.

## Data availability

All data underpinning this review are provided in the main manuscript and supplementary files.

---

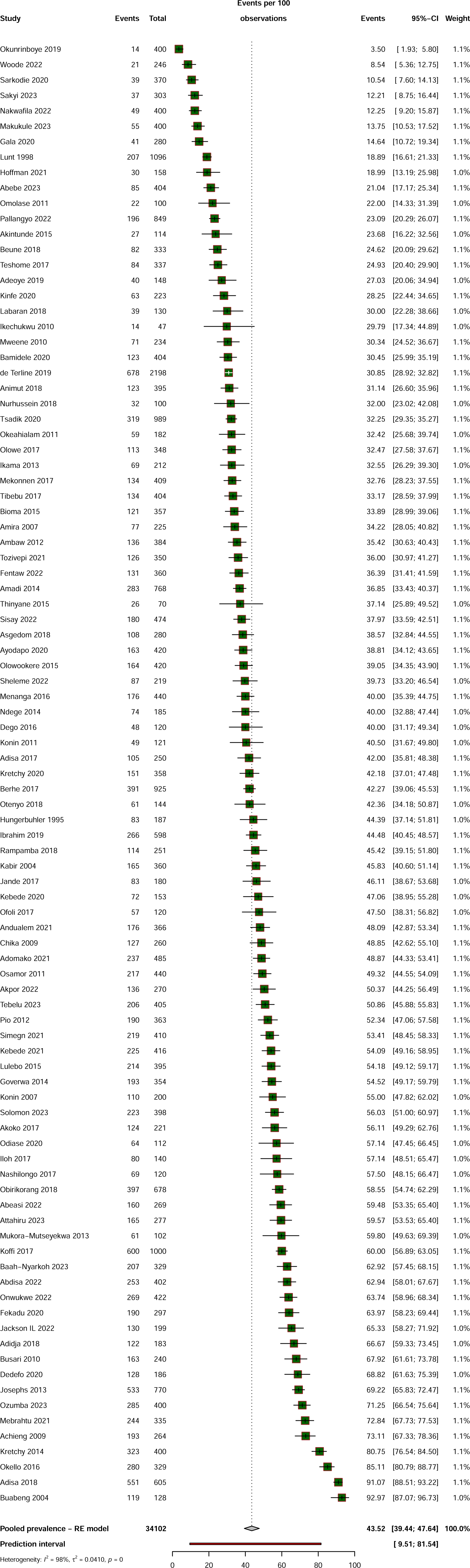

